# A Scoping Review of Algorithmic Equity, Data Diversity, and Inclusive Design in the Transformer Era of Clinical NLP

**DOI:** 10.1101/2025.10.22.25338548

**Authors:** Sheri Grach, Abeer Badawi, Farah Ahmad, Elham Dolatabadi

**Author notes:** These authors contributed equally to this work (Sheri Grach, Abeer Badawi).

## Abstract

The rapid digitization of healthcare has positioned transformer-based natural language processing (NLP) models as powerful tools for managing clinical textual data. Yet their integration into practice raises unresolved questions of equity and inclusivity criteria. This scoping review synthesizes 56 studies published between 2017 and 2024 to evaluate how equity is addressed across three dimensions: algorithmic equity, data diversity and representativeness, and participatory design. Guided by intersectionality and the Digital Health Equity framework, our analysis shows that most equity audits are post hoc and fragmented, with limited impact on model development. Persistent underrepresentation of linguistic, demographic, and clinical subgroups creates what we define as Data Diversity Debt—a structural liability that compounds over time. Participatory design was observed in only 11% of studies, revealing a critical gap in stakeholder inclusion beyond clinicians. Fairness metrics were inconsistently defined, impeding comparability and accountability. To convert descriptive audits into structural repair, we translate these findings into an equity-by-design roadmap that embeds fairness, inclusivity, and accountability across the full lifecycle of healthcare NLP systems. We conclude that equity must shift from reactive auditing to equity-by-design, integrating participatory governance, fairness-aware training objectives, and continuous monitoring to retire Data Diversity Debt and ensure that clinical NLP systems advance rather than reproduce health disparities.

## 1 Introduction

Healthcare delivery is undergoing rapid digital transformation, with new technologies consistently emerging and promising to improve efficiency, accuracy, and patient-centered care^1–5^. Within this landscape, transformer architectures, most notably Bidirectional Encoder Representations from Transformers (BERT), Text-to-Text Transfer Transformer (T5), and Generative Pre-Trained Transformers (GPT), have become the dominant models for clinical natural language processing (NLP)^6–13^. Their technical capabilities, including context-rich embeddings, long-range dependency handling, and large-scale pretraining, have enabled progress in tasks from clinical summarization to diagnostic support^14–17^.

Yet these same models also amplify *equity* risks^18,19^. Empirical evaluations show that transformer outputs can diverge substantially across demographic groups, with serious implications for diagnosis and treatment^20–24^. For example, in a large-scale evaluation of GPT-4 across multiple medical specialties, the model disproportionately described a Black woman in 81% of sarcoidosis vignettes, far exceeding known epidemiology, and correctly prioritized diagnoses in only 37% of cases when race or gender descriptors were altered^23^. Similarly, an analysis of BioBERT revealed strong ethnicity-linked biases, scoring 0.844 (*p* = 0.004) and 0.868 (*p <* 0.001) on benchmarks of hereditary and prevalence-based conditions across different ethnic groups^20^. These examples illustrate that algorithmic inequities are not hypothetical: they are measurable, recurring, and consequential in healthcare contexts.

Despite growing recognition of such risks, scholarship remains fragmented^25,26^. Language bias is a critical but underexplored challenge: most research focuses on English-language models^27^, leaving performance in culturally and linguistically diverse populations insufficiently examined. While multilingual models such as ArabicBERT, IndoBERT, and multilingual BERT have been developed^28^, their capacity to ensure linguistic fairness remains largely untested. Fairness metrics also lack standardization, with inconsistent definitions and applications across studies^29^, complicating cross-study comparison. Most notably, *participatory design*, the involvement of patients, clinicians, and communities in shaping models, remains strikingly under-represented^30^.

This scoping review addresses that gap. Guided by the question: *How are equity-related concerns, including algorithmic equity, data diversity and representativeness, and participatory and inclusive design, addressed in transformer-based NLP for healthcare?* we synthesise evidence from 56 primary studies published between 2017 and 2024. Our analysis is structured around three interconnected equity dimensions:

**Algorithmic equity** involves identification, evaluation, and mitigation of systematic disparities in model performance, with the goal of achieving equitable outcomes across demographic, linguistic, and clinical subgroups^31,32^.

**Data diversity and representativeness** includes breadth, inclusivity, and contextual relevance of datasets used in training and evaluation, spanning demographic, linguistic, geographic, and clinical coverage^33^.

**Participatory and inclusive design** concerns intentional and sustained engagement of patients, clinicians, and community members in developing, evaluating, and deploying NLP models, ensuring alignment with real-world needs and ethical priorities^34,35^.

This review advances the field in three ways. First, it applies *intersectionality*^36^ and the *Digital Health Equity* (DHE) framework^37^ to integrate technical and structural perspectives on equity. Second, it introduces and defines *Data Diversity Debt* as the equity liability that arises when persistent representational gaps compound over time, becoming challenging and costly to correct once models are deployed. Third, it provides the first systematic evaluation of how participatory design is (or is not) embedded in transformer-based NLP research for healthcare. The remainder of this paper is organized as follows. Methods detail the scoping approach and inclusion criteria; Results present thematic findings across the three equity dimensions; Discussion interprets the implications and proposes concrete, evidence-based directions for research, practice, and governance.

## 2 Results

### Study Selection

The database search yielded 1,631 articles published between January 2017 and April 2025. The year 2017 was chosen as a starting point, corresponding to the introduction of transformer architectures into NLP. After removing 653 duplicates, 978 unique records were screened at the title and abstract level. These records were screened against the predefined inclusion and exclusion criteria to assess their preliminary relevance. From these, 193 full texts were assessed for eligibility, and 56 studies were included in the final synthesis (Figure 1).

**Figure 1.**
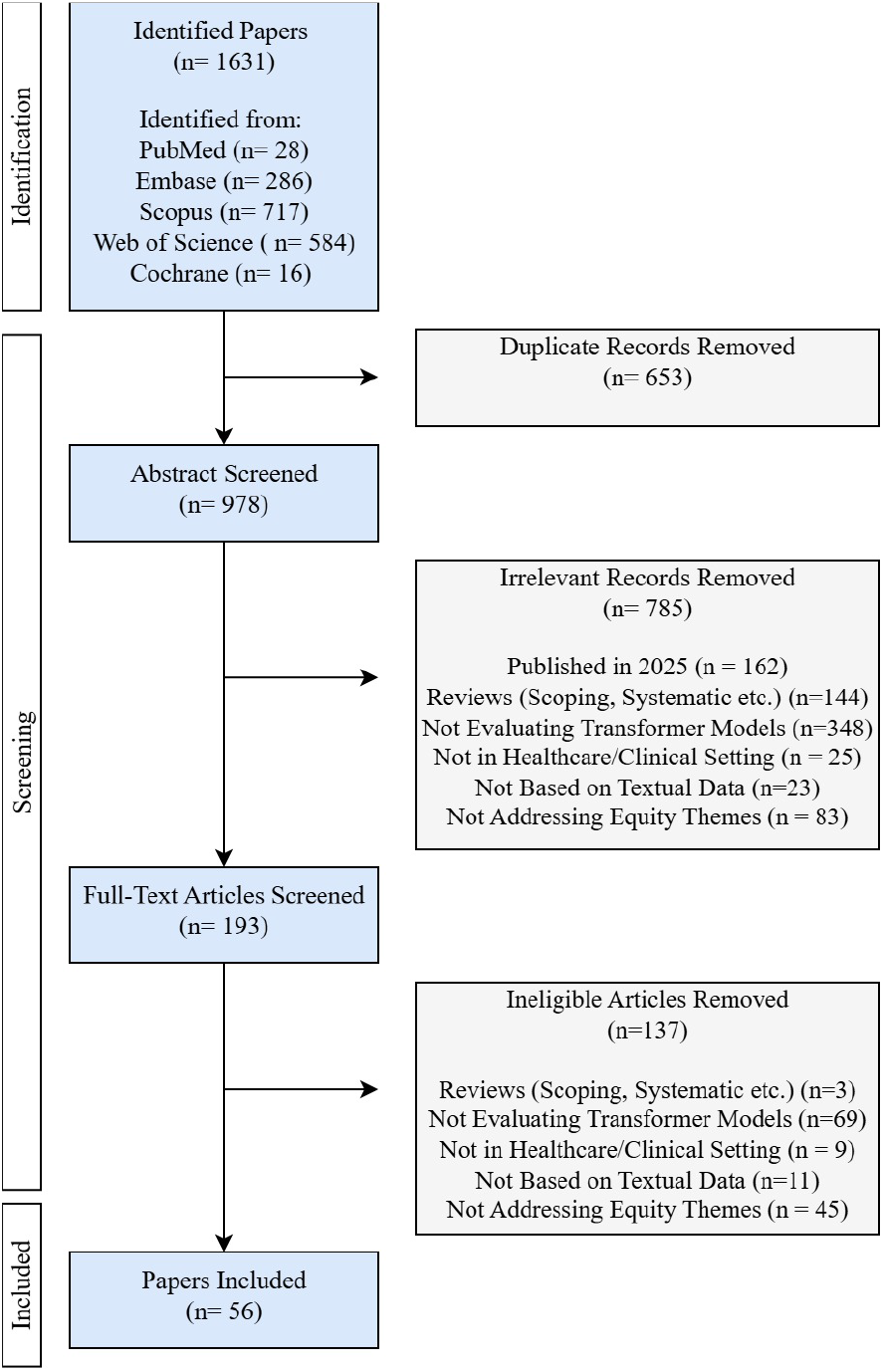
PRISMA-ScR flowchart summarizing the process of study identification, screening, eligibility assessment, and inclusion, with the number of records excluded at each stage.

### Publication Trends

The earliest eligible studies appeared in 2021 (n = 2). Publications increased to 7 in 2022 and 10 in 2023, followed by a sharp rise to 37 studies in 2024, reflecting both the rapid adoption of transformer models in healthcare NLP and the growing salience of equity concerns (Figure 2a). No studies were identified before 2021, consistent with the delayed uptake of transformers in clinical contexts. This trajectory suggests that while equity-focused scholarship is gaining momentum, it remains a nascent field with uneven development. We also analyzed the repeated words from study titles and found that recurring technical and ethical terms are language, clinical, health, bias, ChatGPT, and fairness. While the presence of “bias” and “fairness” signals rising awareness of equity, notable absences include “participatory,” “community,” and “co-design.” This lexical gap suggests that inclusive, stakeholder-driven design remains largely underexplored in transformer-based clinical NLP research, positioning it as a critical blind spot.

**Figure 2.**
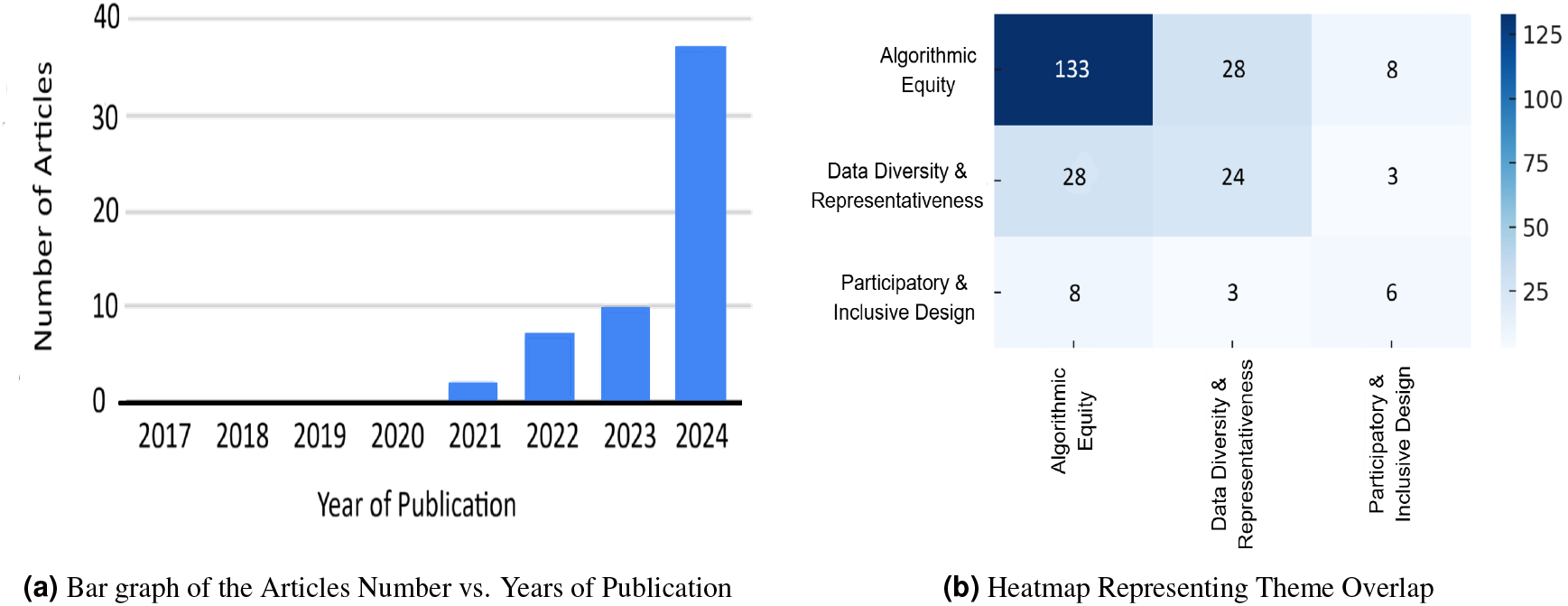
Publication Year and Theme Representation.

A co-occurrence analysis (Figure 2b) revealed that algorithmic equity and data diversity often appeared together (n = 28), underscoring the recognition that biased outcomes often stem from imbalanced training data. In contrast, participatory design showed minimal overlap with the other two themes (n = 8 with algorithmic equity, n = 3 with data diversity), suggesting that social and structural considerations are siloed from technical evaluations. The results reveal a research landscape that is technically sophisticated but socially narrow. While audits of model fairness are increasingly common, systemic equity remains under-addressed due to persistent Data Diversity Debt and the near-absence of participatory design. This disconnect highlights an urgent need to integrate equity across the full model lifecycle, moving beyond technical audits toward inclusive and context-sensitive design.

### Dataset Accessibility and Clinical Domain Distributions

As shown in Figure 3, most studies leveraged private datasets (46.4%), followed by mixed-access datasets combining public and institutional sources (33.9%), and a smaller share of fully public data (19.6%). This distribution indicates a challenge in achieving transparent research, as private datasets often limit external validations. Algorithmic equity analyses were dominant across all data accessibility types, while data diversity and representativeness analyses were less frequent and often clustered around private or mixed datasets. Participatory and stakeholder-driven design was rarely observed, appearing only in scattered instances within public datasets within the general and pediatric domains. Clinical domains were primarily concentrated in general medicine, mental health, and cardiology, revealing the prevalence of textual data in these healthcare domains and the strong presence of transformer-based NLP in patient communication and EHR analysis. In contrast, highly specialized fields like rheumatology, nephrology, and radiology showed very low representation, suggesting limited cross-domain diffusion of transformer applications.

**Figure 3.**
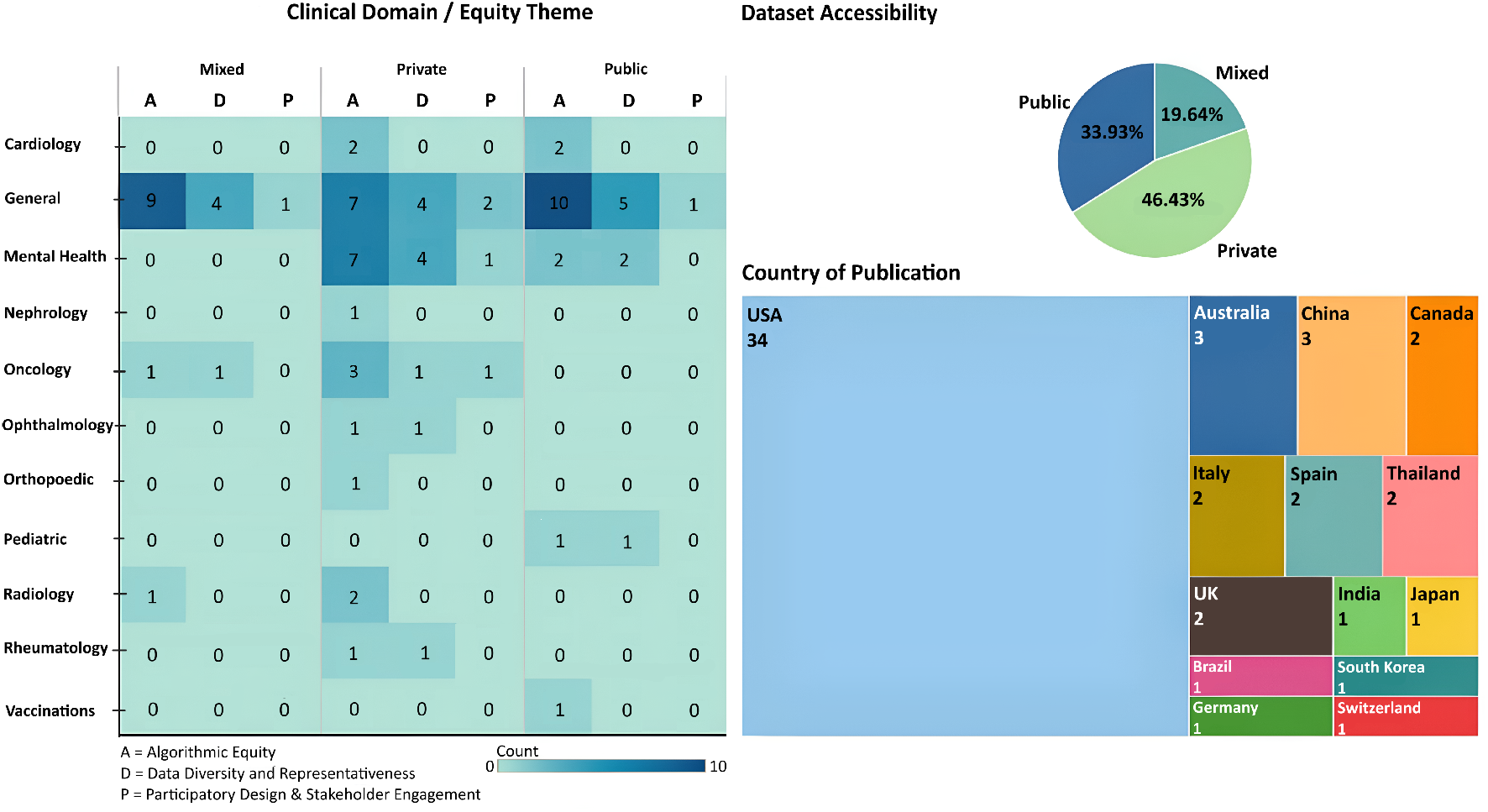
Summary of Dataset Accessibility, Clinical Domains, and Countries of Publication.

### Geographic Distribution of Publications

Country-wise distribution (Figure 3) revealed a strong U.S research dominance (n=34). This was followed by moderate representation from China (n=3) and Australia (n=3). Other countries, including Canada, the U.K., Italy, Spain, and Thailand, each accounted for two studies, while the remaining countries contributed only one study each. The concentration of publications within a small number of countries indicates that research on transformer-based healthcare NLP remains geographically uneven, with limited contributions from other regions.

### Thematic Analysis of Equity Dimensions

Across the 56 included studies, three dimensions of equity were examined with striking imbalance (Figure 4). Algorithmic equity was the most frequently addressed (93%), with studies primarily auditing performance disparities and introducing technical bias-mitigation strategies. Next, data diversity and representativeness were reported in 43% of studies, typically in relation to language coverage, demographic attributes, or clinical context. Yet representation gaps remain persistent, a pattern we term Data Diversity Debt, whereby underrepresentation accumulates over time and amplifies downstream inequities. Participatory and inclusive design was rarely considered (11%, n = 6). Where present, engagement was limited to clinician-in-the-loop validation, with minimal involvement of patients or communities.

**Figure 4.**
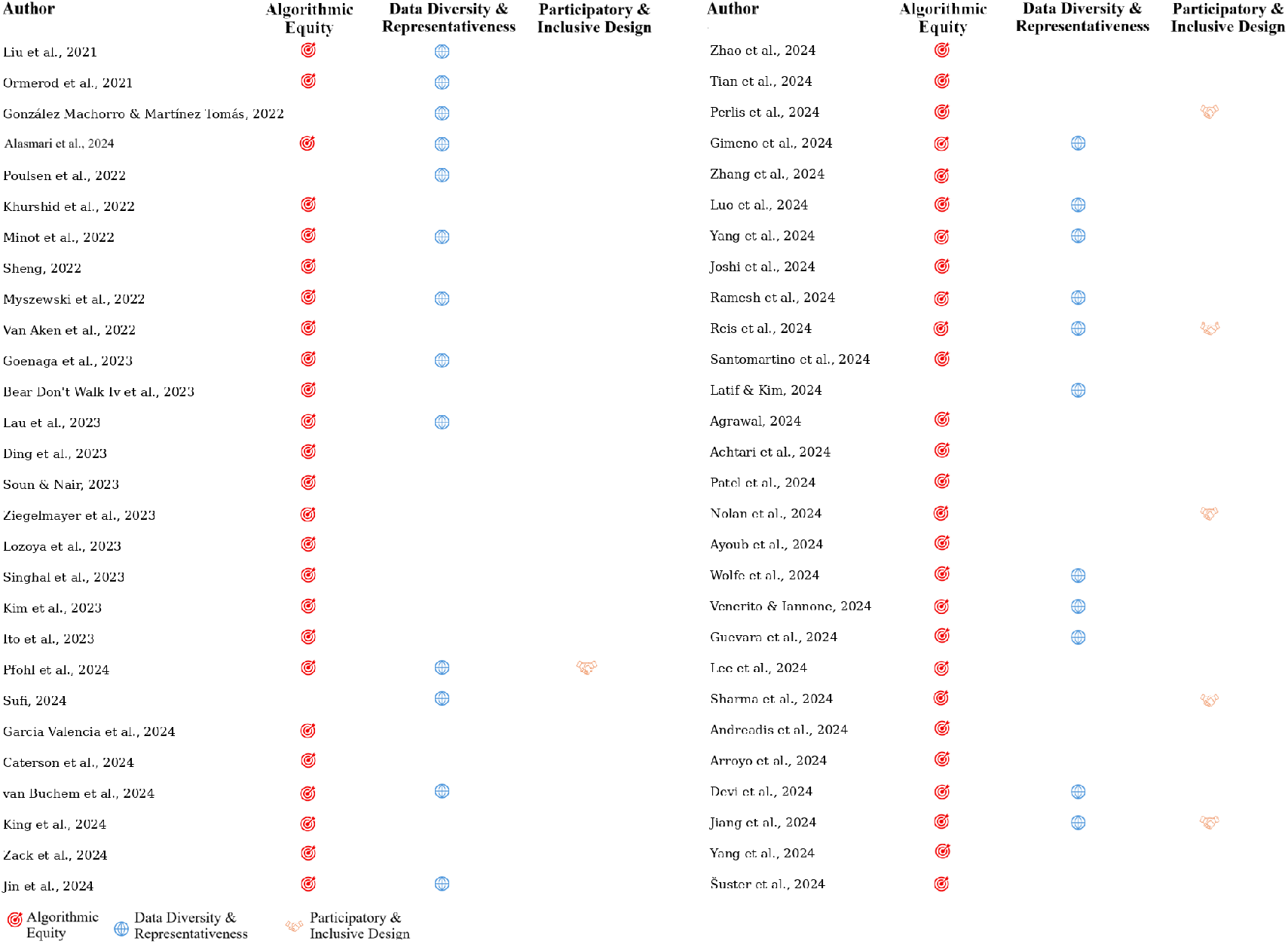
Thematic Summary of Included Studies.

### Algorithmic Equity

Algorithmic equity was the most frequently examined theme, appearing in 93% of included studies as shown in Figure 4. Evidence shows that transformer-based NLP models, reliant on large-scale pretraining^6,9^, contextual embeddings, and attention-weighted representations, consistently encode representational and contextual biases that translate into disparities across demographic, linguistic, and clinical subgroups. Unlike traditional classifiers, transformers affect not only predictive accuracy but also style, tone, and empathy, highlighting their structural vulnerability to inequity.

In clinical documentation, GPT-3 and ChatGPT produced inappropriate or culturally inconsistent phrasing, omissions, and insertions, embedding bias in generated text^38^. Semantic similarity evaluations likewise reinforced skewed associations^39^. Large generative models, including GPT-4 and Med-PaLM 2, perpetuated racial, gender, and socioeconomic stereotypes during question-answering tasks^23,40^, while over 1,000 GPT-4 simulations revealed systematic in-group favoritism across demographic and social categories^41^. These findings suggest that stereotypes are deeply embedded within transformer contextual representations. Auditing methods also showed limitations. Zero- and few-shot prompting produced weak and inconsistent estimates of bias in clinical trial evaluations^42^, reflecting transformer-specific instability where outputs are highly sensitive to prompt phrasing. This methodological fragility raises concerns about the reliability of widely used audit pipelines.

Linguistic disparities were repeatedly observed. GPT-4 discharge instructions were more accurate in English than Spanish^43^, and CDC vaccine guidelines were more faithfully replicated in English^44^. English-trained models underperformed when adapted to Spanish disability annotation^45^, and kidney transplant FAQs translated with GPT-3.5 and GPT-4 contained subtle inconsistencies^46^. Fine-tuned AraBERT on Arabic medical text significantly improved on question-answering^47^, highlighting the importance of language-specific adaptation. Demographic inequities were also prevalent. GPT-4 misclassified minority patients in simulated triage^48^, recommended poorer psychiatric treatments for Black women compared to White men^49^, and produced more severe or suboptimal diagnoses for Black and Asian patients^50^. In radiology, models exhibited age-related disparities^51,52^, while EMR-based tasks showed gender- and age-related bias that required debiasing^53^. Even when overall diagnostic accuracy was stable, subgroup-level differences emerged in urgent care recommendations^54^, revealing what we term latent attention-driven disparities.

Bias in interpersonal outputs was equally noticeable. ChatGPT’s empathy varied by patient race and gender in oncology queries^55^; sentiment bias appeared in mental health contexts^56^; GPT-3 produced gendered language emphasizing emotionality for women and competence for men^57^; and GPT cardiovascular risk assessments shifted by patient sex^58^. These inequities shape not only accuracy but also trust and engagement. Equity concerns were most consequential in high-risk mental health settings. GPT-4 matched clinicians overall in suicidal ideation prediction but showed inequitable subgroup performance^59^. In evaluation scenarios, LLM-as-judge evaluations systematically diverged from human clinician ratings in mental health dialogue assessment, revealing inflated scores and hidden biases in fairness and safety^60^. In Brazil, GPT-4 and LLaMA 3 medication instructions improved with prompt engineering to reduce gender bias^61^, while oncology comparisons revealed persistent race- and gender-based disparities across LLMs^55^. These findings show that while targeted technical interventions show promise in reducing inequities, their adoption remains inconsistent.

Mitigation strategies included synthetic data generation, which enriched underrepresented subgroups and reduced disparities^62^, GAN-enhanced BioBERT for adverse event detection^63^, and privacy-preserving methods that inadvertently introduced new demographic gaps^64^. Open-weight models such as LLaMA and Mistral, when fine-tuned on curated datasets, achieved equitable performance in resource-limited settings^65^. Design-level interventions such as attention regularization, gender-term masking^66^, cohort expansion pipelines^67^, and fairness assessments in SDOH extraction^68^ demonstrated potential, while prompt phrasing sensitivity remained a key determinant of equity outcomes^69^. Fairness audits are increasingly integrated into radiology pipelines^70^, though masked language models such as BioBERT and PubMedBERT continue to reveal demographic biases^71^, and race/ethnicity extraction models sometimes rely on stigmatizing descriptors^72^. In summary, algorithmic equity in healthcare transformers is compromised by biases built into their design, training data, and text-generation processes. The reviewed studies highlight the urgent need for standardized fairness metrics, robust multi-stage audits, and integration of sociocultural perspectives to ensure these models do not entrench disparities but advance equitable care.

### Data Diversity and Representativeness

Data diversity and representativeness were reported as persistent challenges, with inequities emerging across demographic, linguistic, geographic, and clinical dimensions. Because transformer embeddings reflect the distribution of their pretraining corpora, underrepresentation in training data directly propagates into biased predictions and reduced generalizability^57,62^. Synthetic augmentation was widely explored as a mitigation pathway. GPT-generated clinical data enriched representation for low-resource groups^62^, improved preventive care classification^73^, and enhanced risk modeling with BART and T5^73^. Yet, synthetic notes also reproduced gender stereotypes^57^, reflecting a transformer-specific tension: augmentation expands coverage but also mirrors corpus biases. Open-weight models such as LLaMA and Mistral achieved equitable summarization when fine-tuned on curated data, demonstrating the value of deliberate dataset curation in resource-limited contexts^65^.

Parameter-efficient strategies highlighted adaptability under scarcity. Prefix-tuning accurately estimated depression severity with limited transcripts^74^, while multilingual cognitive-impairment detection varied with dataset richness^75^. For rare conditions, ClinicalBERT and BioBERT outperformed structured fields in opioid use disorder detection but remained constrained by annotation gaps^76^. Linguistic and cultural representativeness emerged as a key factor. Monolingual BETO outperformed mBERT and XLM-R for Spanish disability annotation^45^; GPT-based Spanish translations of kidney FAQs were syntactically accurate but incomplete^46^; and Arabic models trained on medical Arabic datasets substantially outperformed the hybrid models^47^. Region-specific models such as CHIMED-GPT and ChatZOC achieved superior accuracy and expert alignment compared to general-purpose LLMs^77^. Conversely, GPT-4 was consistently more accurate in English than in non-English healthcare queries^78^.

The structural fragmentation of healthcare data also limited equitable performance. Hybrid BERT–TabNet integration improved psychiatric rehabilitation predictions by bridging structured and unstructured sources^79^, while adversarial training and attention regularization produced gender-sensitive embeddings in ICU readmission tasks^66^. Large-scale EHR analyses confirmed that demographic underrepresentation drove diagnostic disparities^53^. In Brazil’s e-prescription system, prompt-engineered GPT instructions reduced gender bias and improved personalization^61^. Dataset provenance and confounder control further affected representativeness. Provenance adjustments improved model transferability across institutions^80^, while sentiment analysis of fibromyalgia narratives demonstrated transformers’ utility in noisy, low-resource domains^81^. GPT-4 also improved eligibility classification for clinical trials, reducing manual adjudication bias^82^.

Taken together, the studies demonstrate that transformers’ performance and fairness are inseparable from the inclusivity of their training data. While synthetic augmentation, multilingual adaptation, and fairness-aware modeling offer partial remedies, upstream curation that is demographically balanced, linguistically diverse, and contextually grounded remains essential to prevent persistent disparities in clinical applications.

### Participatory Design and Stakeholder Engagement

Participatory and inclusive design was the least represented theme, appearing in only 11% of studies as shown in Figure 4. Yet this gap is striking given that transformers, with generative and prompt-sensitive outputs, risk producing clinically plausible but socially misaligned responses without stakeholder oversight. The most comprehensive example was an equity-focused harm assessment toolbox for GPT-4 and Med-PaLM 2, co-created with clinicians and community advisors, which revealed hidden biases and provided a replicable evaluation framework^40^. Other studies involved clinicians in benchmarking diagnostic outputs^54^ or assessing GPT-based Spanish kidney transplant translations^46^, revealing gaps invisible to automated checks. In some contexts, participatory feedback directly improved outputs. Brazilian pharmacists and physicians iteratively refined GPT-generated medication instructions, reducing gender bias and improving personalization^61^, while radiology reviewers applied fairness and coherence criteria to guide refinements in imaging impressions^70^.

Similar expert-in-the-loop frameworks were used in suicide risk prediction^59^, social determinants extraction^83^, and oncology trials^84^, where clinician adjudication shaped transformer evaluation. Specialized engagement also informed model alignment. For example, simulated patient value profiles tested surrogate decision-making in GPT-3.5 and BERT^85^, while clinician input during hybrid BERT–TabNet development improved psychiatric rehabilitation predictions^79^. In the mental health domain,^86^proposed the SAFE-i and HAAS-e frameworks, embedding participatory, human-centered evaluation and co-creation to align LLMs with ethical and clinical equity needs in healthcare. Despite these examples, participation remained almost entirely limited to technical and clinical experts, with no documented involvement of patients, caregivers, or communities. Absent mechanisms such as patient advisory boards or collaborative annotation highlight a major equity gap. Given transformers’ sensitivity to social framing and narrative diversity, expanding engagement to marginalized and underserved groups is essential for contextual alignment, trust, and equitable deployment.

## 3 Discussion

The rapid rise of transformer-based NLP models in healthcare brings significant promise, yet our scoping review of 56 studies published between 2017 and 2024 reveals a critical imbalance in how equity is addressed. Across the 56 studies, three patterns dominate: (i) algorithmic equity is examined in 93% of papers, typically via subgroup audits; (ii) data diversity and representativeness appear in 43%, often as reactive fixes; and (iii) participatory and inclusive design is rare (11%, *n* = 6). Together with the co-occurrence results (Algorithmic Equity ↔ Data Diversity: 28 studies; Participatory Design ↔ Algorithmic Equity: 8; Participatory Design ↔ Data Diversity: 3; Fig. 2b), these findings indicate a structural imbalance: the field measures inequities downstream but underinvests in upstream representation and stakeholder governance.

Most equity evaluations are post hoc, conducted after model training, limiting their ability to alter datasets, objectives, or pretraining corpora. Empirically, disparities surface across clinical and biomedical NLP tasks: large language models such as GPT-4 have shown unequal error rates in synthetic triage simulations for minority patients^48^ and lower-quality treatment recommendations for marginalized groups^49^. Audit instability has also been observed,few and zero-shot evaluations often produce inconsistent bias scores across prompt templates or sampling seeds, echoing broader findings that in-context learning in transformers is highly sensitive to prompt phrasing and position^87,88^. Such sensitivity arises from the self-attention mechanism’s reliance on contextual token weighting^89^ and from differences in pretraining objectives, masked-language modeling (e.g., BERT, BioClinicalBERT) versus causal autoregressive modeling (e.g., GPT-style), which shape downstream inductive biases^90^. Additionally, decoding strategy (e.g., greedy vs. nucleus sampling) affects both factual accuracy and response tone^91,92^. Framing equity audits as a “final check”, therefore, limits their corrective leverage over the architectural and training choices that ultimately govern transformer behavior.

The strong co-occurrence between algorithmic equity and data diversity (28 studies) indicates a shared recognition that representational gaps are root causes of inequity. Yet mitigation efforts remain fragmented. Synthetic data augmentation can improve minority-class signal but may also amplify stereotypical associations; multilingual adaptation (e.g., AraBERT, XLM-R) improves dialect-specific performance yet highlights the structural limits of English-dominant pretraining; open-weight transformers fine-tuned on curated local corpora can reduce disparities but rely heavily on institutional curation capacity. Mechanistically, disparities across languages and subpopulations are consistent with transformer-specific factors: subword tokenizers (e.g., BPE, WordPiece) tend to over-fragment low-resource languages or dialectal variants, inflating sequence length and degrading representational coherence^93^; pretraining corpus imbalance, where English text dominates, biases contextual embeddings and cross-lingual alignment, limiting transfer to underrepresented languages^94^; and context-length constraints in transformer architectures (finite positional embeddings or memory windows) can truncate long clinical narratives, increasing the risk of discarding salient or demographically specific information^95,96^. We frame this as *data diversity debt*, the equity liability that arises when populations, languages, regions, or care settings are systematically underrepresented in the datasets used to train clinical NLP models, becoming harder to remediate once embedded in large pretrained models. In this framing, inclusivity is not a dataset attribute but a structural determinant of model equity.

Only six studies incorporated stakeholder engagement, typically clinicians validating outputs, with scarce involvement of patients, caregivers, or community representatives. Where engagement occurred, it was consequential: Spanish-speaking nephrologists surfaced completeness and nuance issues in GPT-based transplant frequently asked questions translations^46^; a participatory harm-assessment toolbox for GPT-4/Med-PaLM 2 uncovered hidden failure modes and offered a replicable evaluation template^40^. Conversely, high-stakes settings such as suicide risk prediction showed subgroup variability^59^ without embedded participatory oversight to anticipate or monitor harms. In instruction-tuned transformers, participatory oversight is uniquely consequential: prompt templates, refusal policies, readability targets, and culturally sensitive phrasing all influence how responses are generated. Without such governance, models may achieve strong benchmark performance yet remain misaligned with the linguistic, cultural, and relational realities of care^97^.

Findings across themes and co-occurrences support a systemic view that unrepresentative data generate biased embeddings^98^; weak, late-stage audits miss or under-correct harms; absent participatory governance leaves context-specific risks undetected. The intersectionality lens explains compounding disadvantages across demographics and languages^36^, while the DHE framework situates these model-level issues within infrastructural access, digital literacy, and institutional trust^37^. Equity, therefore, cannot be added to one stage; it must be embedded across the lifecycle.

### Guiding Principles

The acceleration of publications since 2021, peaking in 2024, confirms that equity in transformer-based clinical NLP has moved to the mainstream. To convert descriptive audits into structural repair, we translate these findings into an equity-by-design roadmap with three integrated implications for research and governance (Figure 5).

**Figure 5.**
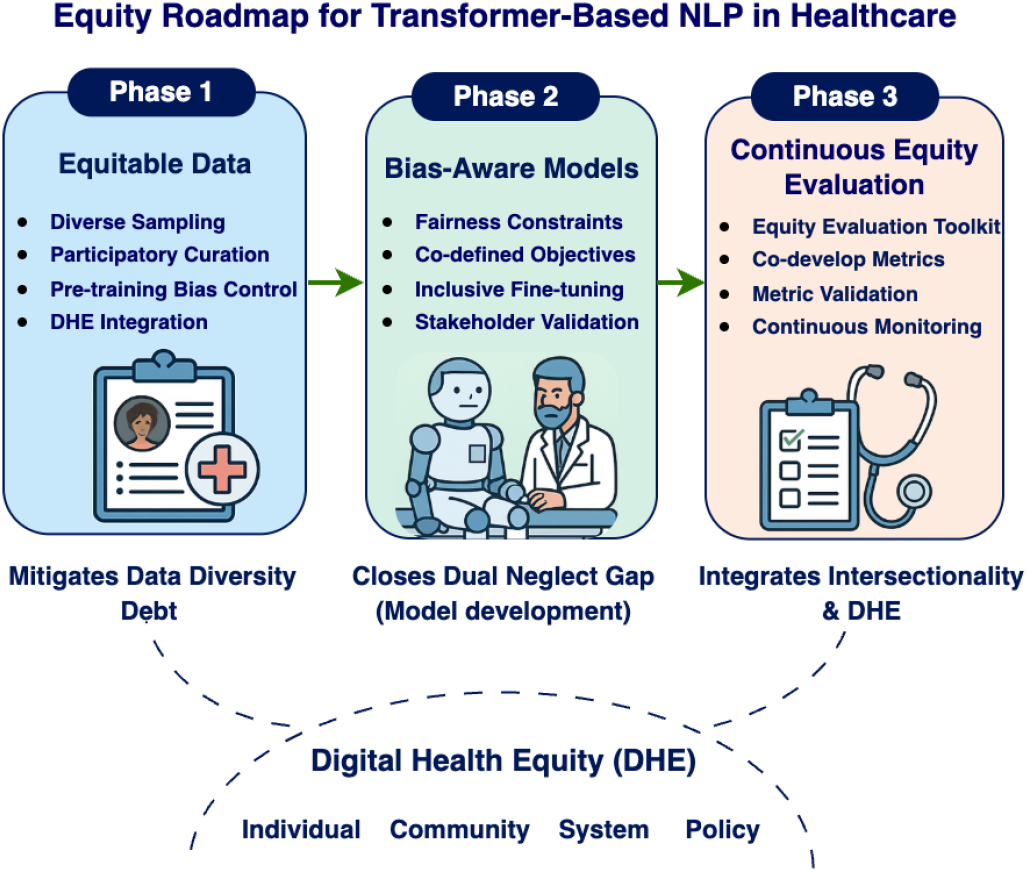
Equity Roadmap for Transformer-Based Healthcare NLP.

#### Phase 1. Retire “data diversity debt” through upstream curation

Given the tight coupling we observed between performance disparities and representational gaps, inclusive data practices must become first-order infrastructure. Concretely, datasets should (i) intentionally sample underrepresented dialects, demographics, regions, languages, and clinical settings with transparent composition reporting; (ii) use participatory curation—patient and caregiver advisory boards, community workshops, and compensated annotation—to align corpora with lived care contexts; (iii) apply pre-training bias controls (balancing, targeted augmentation, metadata tagging) before any model fitting; and (iv) incorporate DHE factors (infrastructure access, digital literacy, trust) to avoid reifying structural barriers at the point of data collection. These measures directly address the upstream drivers of the disparities repeatedly documented in our corpus.

#### Phase 2. Bake fairness into model objectives, not just evaluations

Because audits alone did not prevent recurrent inequities across triage, radiology, psychiatry, oncology, and multilingual tasks, fairness constraints must enter at training time. Implications include: (i) participatory design expansion—stakeholders help define fairness goals, surface failure modes, and co-specify constraints; (ii) adoption of fairness-aware objectives (adversarial debiasing, representation regularization, constraint optimization) alongside domain- and language-specific fine-tuning for underserved populations; and (iii) prospective tests of design choices on usability, accessibility, and clinical applicability, combining quantitative parity metrics with qualitative stakeholder judgment. This shifts equity from post hoc auditing to preemptive model shaping, consistent with the sensitivity to prompts and context highlighted in our results.

#### Phase 3. Standardize equity evaluation and make it continuously accountable

Unstable few-/zero-shot audits and heterogeneous metrics in the literature argue for a shared toolkit and continuous monitoring. We therefore recommend: (i) an equity-centered evaluation toolkit with templates for stakeholder-involved audits, intersectional subgroup testing, and lifecycle benchmarking; (ii) institutionalization of standardized equity metrics in policy, certification, and procurement (models should not be deployed without documented subgroup performance and uncertainty); (iii) methodological work to identify which metrics best capture technical *and* clinical parity for specific tasks, with open benchmarks and provenance reporting; and (iv) participatory review and real-world monitoring, counterfactual testing, drift detection, and longitudinal harm tracking, to catch delayed or compounding effects after deployment.

Across phases, the field should treat inclusive data infrastructure, fairness-aware training, and participatory oversight as *co-requisites*. Practically, this means: requiring dataset cards with demographic and linguistic coverage; reporting intersectional results alongside aggregate metrics; documenting participatory inputs to design; and making post-deployment equity monitoring a condition of clinical use. Absent these steps, transformer-based NLP risks scaling the very inequities it aims to reduce; with them, the trajectory documented here can pivot from measurement to material improvement in health equity.

These findings highlight the need for a paradigm shift in how equity is conceptualized and operationalized. Equity must be embedded across the model lifecycle as a structural commitment. As illustrated in Figure 5, the Equity Roadmap provides a pathway forward: (1) equitable data practices to address Data Diversity Debt, (2) bias-aware model development, (3) fairness evaluation with intersectional audits and continuous monitoring. For researchers, this means moving beyond simple performance metrics toward equity audits that capture clinical impact. For clinicians, it highlights the importance of establishing mandates, benchmarks, and governance structures that integrate equity into the deployment of NLP models in healthcare. Only through such integrated and anticipatory approaches can transformer-based NLP systems move from technical sophistication to genuine social accountability.

In conclusion, this scoping review underscores that equity in transformer-based healthcare NLP must evolve from isolated audits to a sustained, lifecycle-wide commitment. When we reframe data diversity, algorithmic fairness, and participatory governance as interdependent rather than sequential goals, the field can shift from reactive mitigation to structural inclusion. Embedding equity-by-design principles into data infrastructure, model development, and continuous evaluation—supported by transparent benchmarks and participatory oversight—offers a path toward clinically meaningful and socially accountable AI. Achieving this transformation requires collaboration between researchers, clinicians, policymakers, and communities to ensure that future NLP systems do not merely perform well but also perform justly.

## 4 Methodology

This study used a scoping review methodology to examine equity-related concerns in transformer-based NLP for healthcare. The review focused on three ethical dimensions—algorithmic bias, data diversity, and participatory design—capturing how these issues are addressed and where critical gaps remain. We followed the Arksey and O’Malley framework^99^ and adhered to the PRISMA-ScR checklist^100^ to ensure methodological rigor. This structured process guided inclusion criteria, study selection, data extraction, and synthesis. The approach was chosen to map the breadth of an emerging, interdisciplinary field and to highlight variability in how equity is operationalized in clinical NLP research. Situating transformer models within an equity framework, the review aims to inform the ethical and socially responsible integration of AI into healthcare practice and policy, ensuring that technological advances support rather than undermine health equity across diverse populations.

### Frameworks and Approach

Drawing from critical realism research^101^, this review was guided by the intersectionality approach^36^ alongside the Digital Health Equity (DHE) framework^37^. The intersectionality approach offered a critical lens for examining how overlapping social identities, such as race, gender, and language background, interact with systemic structures to shape the development and outcomes of transformer-based NLP models in healthcare. In parallel, the Digital Health Equity framework offered a structured perspective for assessing how digital determinants of health, such as access, trust, and digital literacy, along with layered contexts spanning the individual, interpersonal, community, and systemic levels, influence the adoption and equitable implementation of NLP systems in clinical settings. These frameworks informed the design of the data extraction template, guided the categorization of studies during screening and charting, and shaped the thematic interpretation of results. We embed these frameworks into the analytic process to move beyond superficial mentions of equity to interrogate how structural and contextual forces shape transformer-model deployment in healthcare. The application of both frameworks enabled a more nuanced and contextually grounded understanding of equity-related concerns in the development and use of transformer-based NLP tools for healthcare.

### Eligibility Criteria

To ensure the relevance and consistency with the research objectives, strict inclusion and exclusion criteria were applied. Studies were eligible for inclusion if they applied transformer-based natural language processing models, such as BERT, T5, GPT, or other domain-specific variants like BioBERT or ClinicalBERT, within healthcare or clinical contexts. Eligible applications included clinical text classification, medical summarization, question answering, triage, and related tasks involving textual healthcare data. In addition to using transformer models, studies had to be meaningfully engaged with at least one equity-related theme. These included: (1) algorithmic equity — encompassing the identification, evaluation, and mitigation of systematic disparities in model performance, as well as the pursuit of equitable outcomes across demographic and clinical subgroups; (2) data diversity and representativeness — addressing the breadth, inclusivity, and contextual relevance of datasets used for training and evaluation, with particular attention to the representation of historically underserved populations; and (3) participatory and inclusive design — involving clinicians, patients, community representatives, or other stakeholders throughout the model lifecycle to ensure contextual alignment, trust, and real-world relevance.

Eligible studies were primary research articles published in peer-reviewed journals or conference proceedings, written in English, and published between January 2017 (the introduction of transformer architectures) and December 2024. Although the database search extended to April 2025, articles published in 2025 were excluded to maintain consistency with the review window. Studies were excluded if they: (i) focused solely on performance metrics (e.g., accuracy, F1) without equity considerations; (ii) used non-transformer models (e.g., traditional machine learning or recurrent neural networks); or (iii) were non-peer-reviewed sources such as opinion pieces. These criteria ensured methodological rigor and substantive engagement with the review’s central themes.

A structured literature search was conducted across five major databases: PubMed, Embase, Scopus, Web of Science, and Cochrane Library. These databases were selected for their broad coverage of biomedical, clinical, and interdisciplinary research domains relevant to the objectives of this scoping review. The search strategy used a combination of key terms and Boolean operators to reflect three main concept clusters: (1) NLP/transformer-based models, (2) equity-related concerns, and healthcare or clinical applications. The final search terms included: (“transformer models” OR “BERT” OR “BioBERT” OR “ClinicalBERT” OR “NLP” OR “LLM” OR “T5” OR “GPT”) AND (“bias” OR “algorithmic bias” OR “equity” OR “intersectional bias” OR “data diversity” OR “data scarcity” OR “participatory design” OR “fairness” OR “ethical AI”) AND (“healthcare” OR “clinical” OR “medical” OR “health” OR “clinic” OR “clinical decision support” OR “electronic health records” OR “patient care”)

### Data Extraction and Analysis

A structured extraction table was developed to systematically chart relevant information from each included study, as shown in Table 1. Key variables extracted included the title, authors, publication year, study design, healthcare context, specific transformer model(s) used (i.e., BERT, T5, BERT variants), dataset type (i.e., clinical notes, EHRs), and the equity dimensions addressed. To align with the review’s guiding frameworks, deductive categories included algorithmic equity, data diversity and scarcity, and participatory design. In addition, findings were categorized according to the levels of the Digital Health Equity (DHE) framework, including individual, interpersonal, community, and societal, to assess how equity-related concerns were situated across different layers of the socio-ecological context. The extracted data were then coded and analyzed using a combined deductive-inductive thematic approach. Predefined codes reflected the equity dimensions, while inductive coding allowed additional themes to emerge from the literature. The intersectionality lens and the DHE framework informed this process, ensuring that coding captured both technical and contextual factors. The final synthesis grouped studies under the three main equity dimensions, providing a structured basis for presenting the results.

**Table 1.**
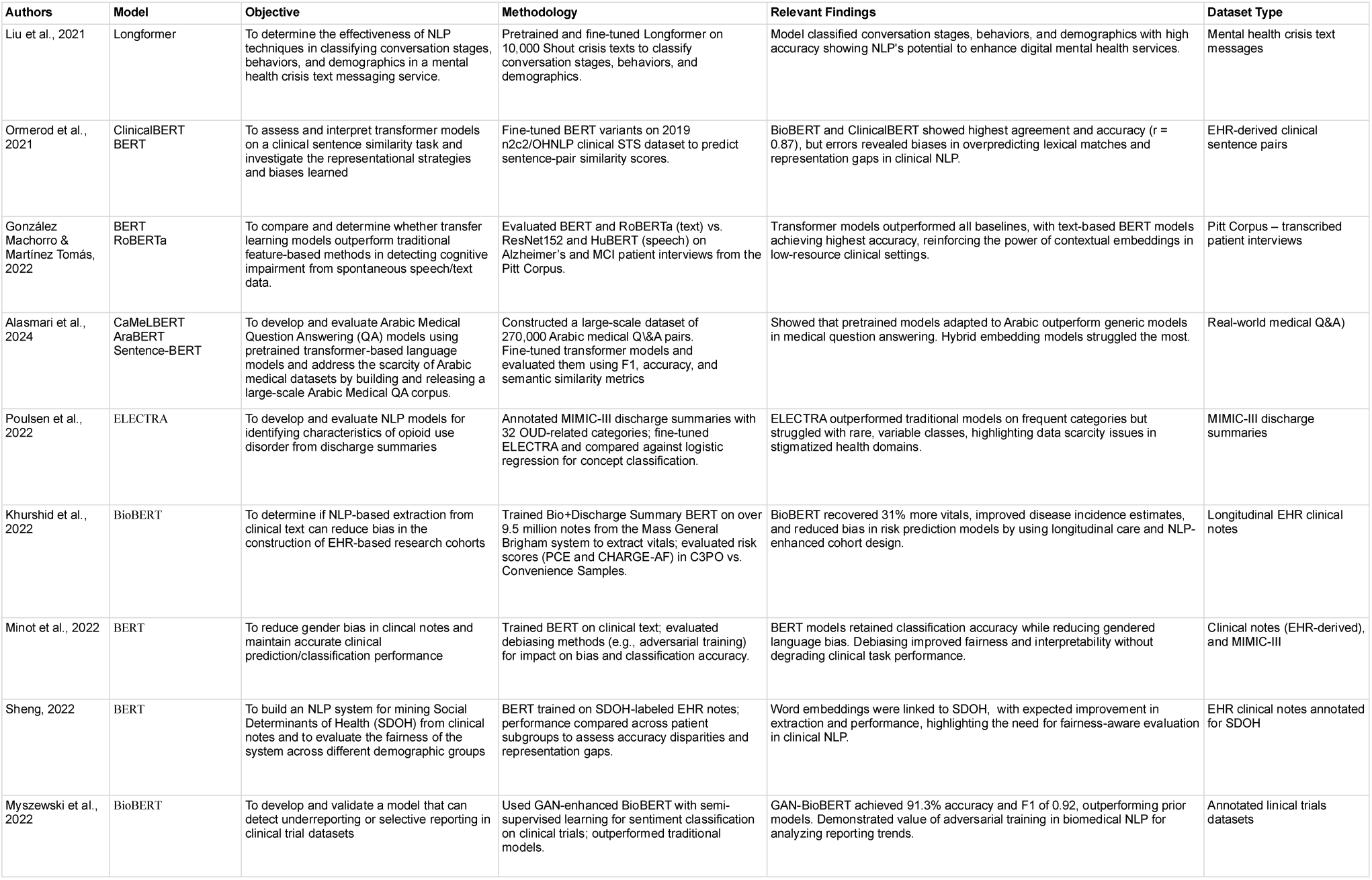
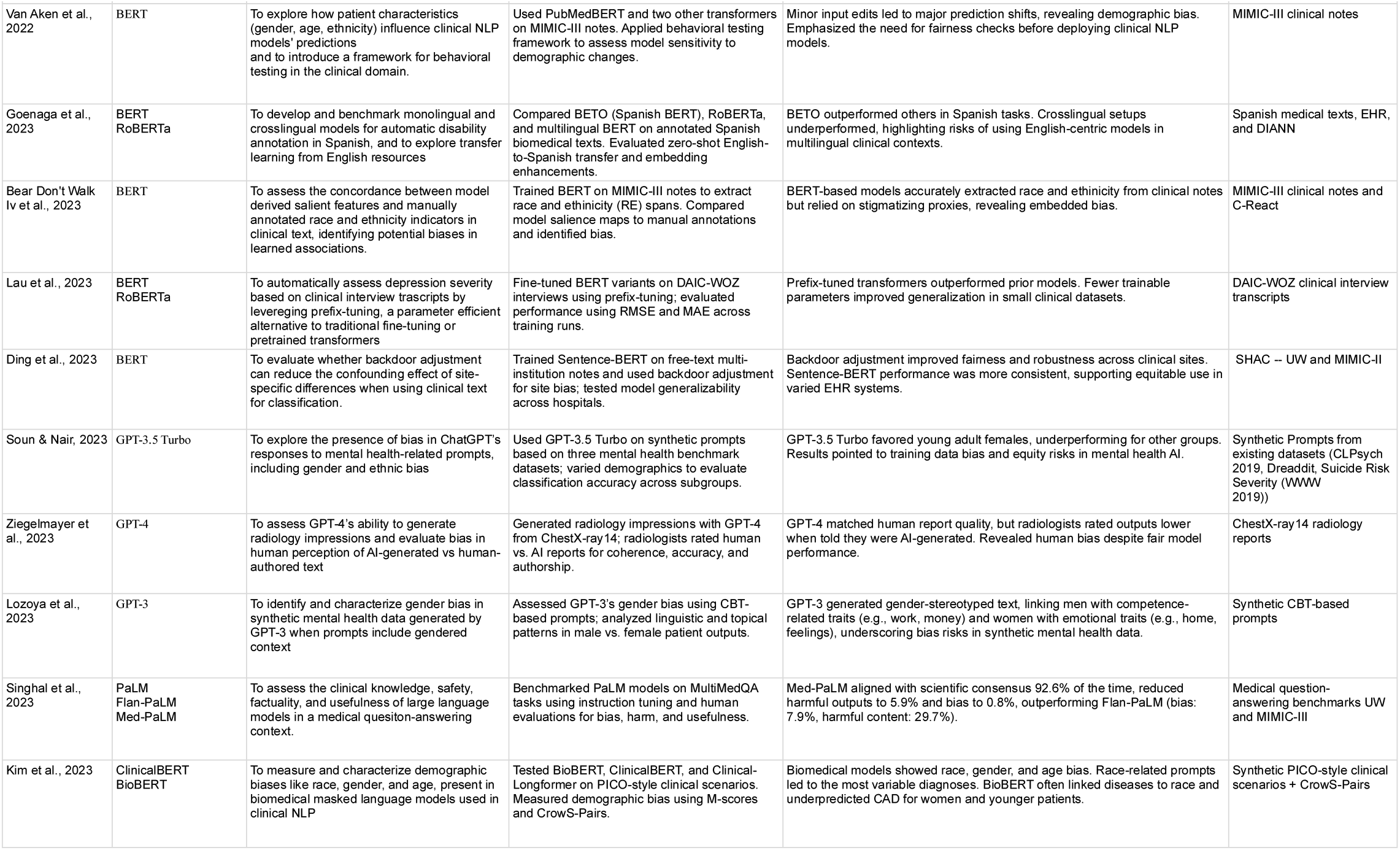
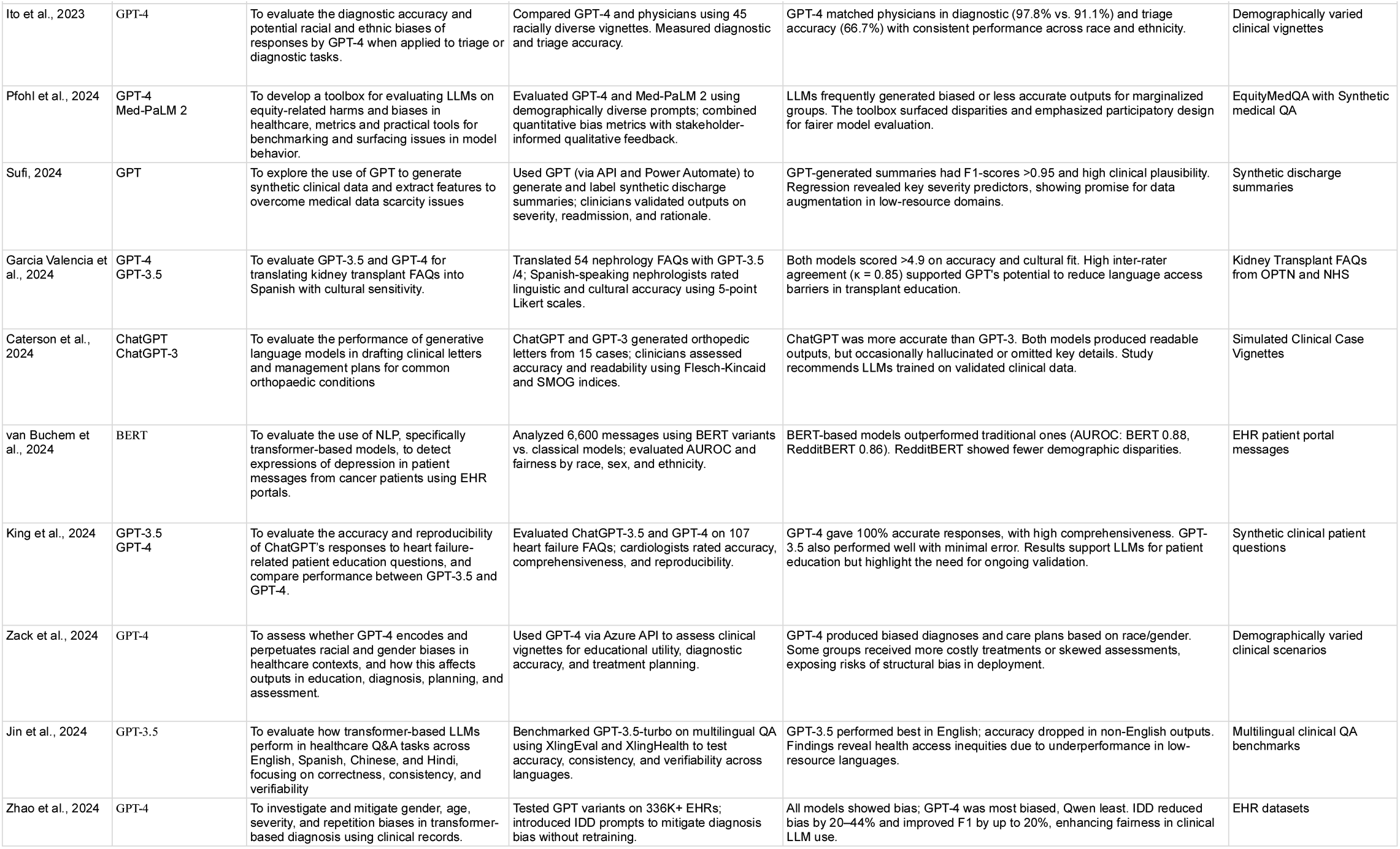
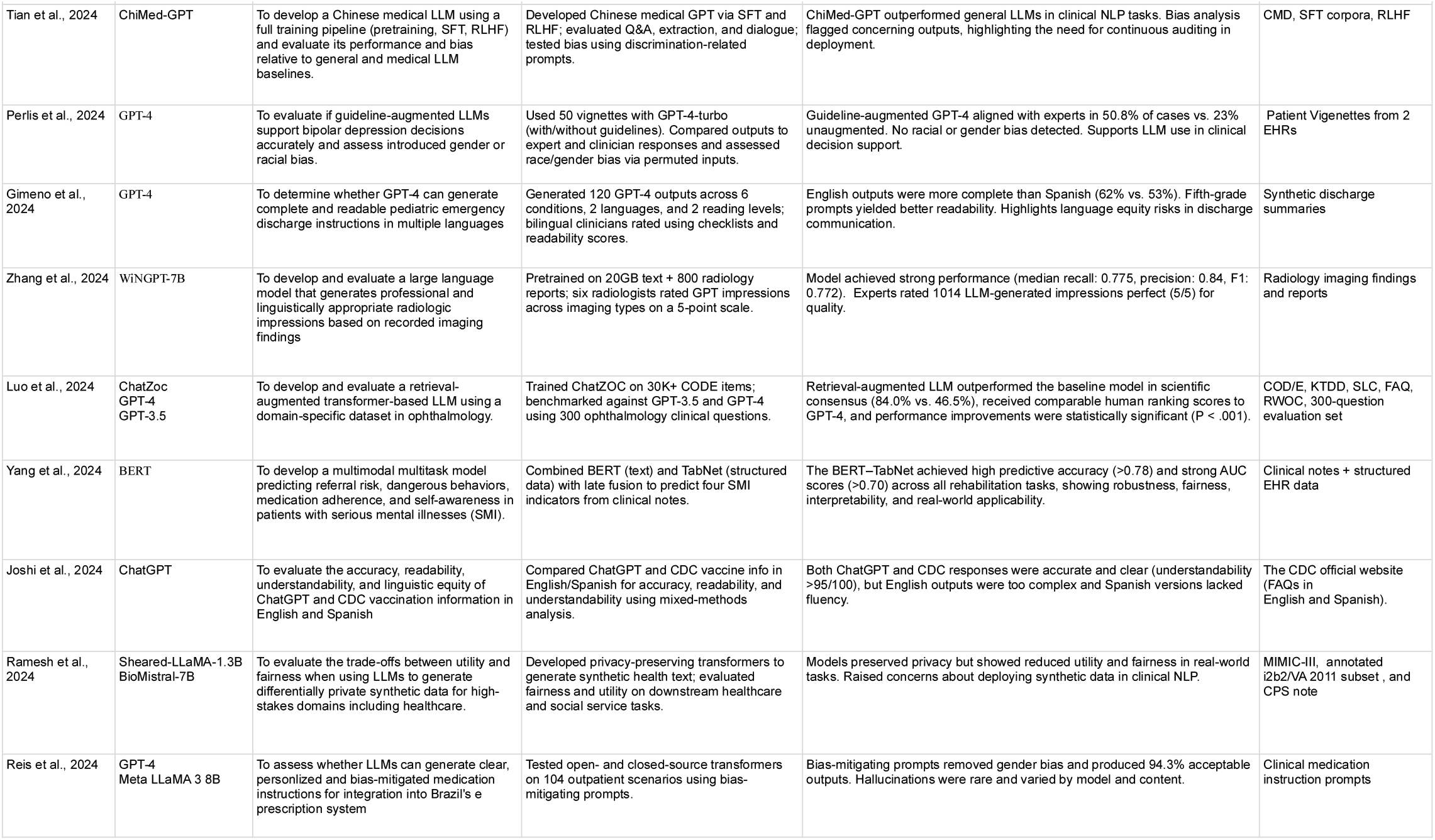
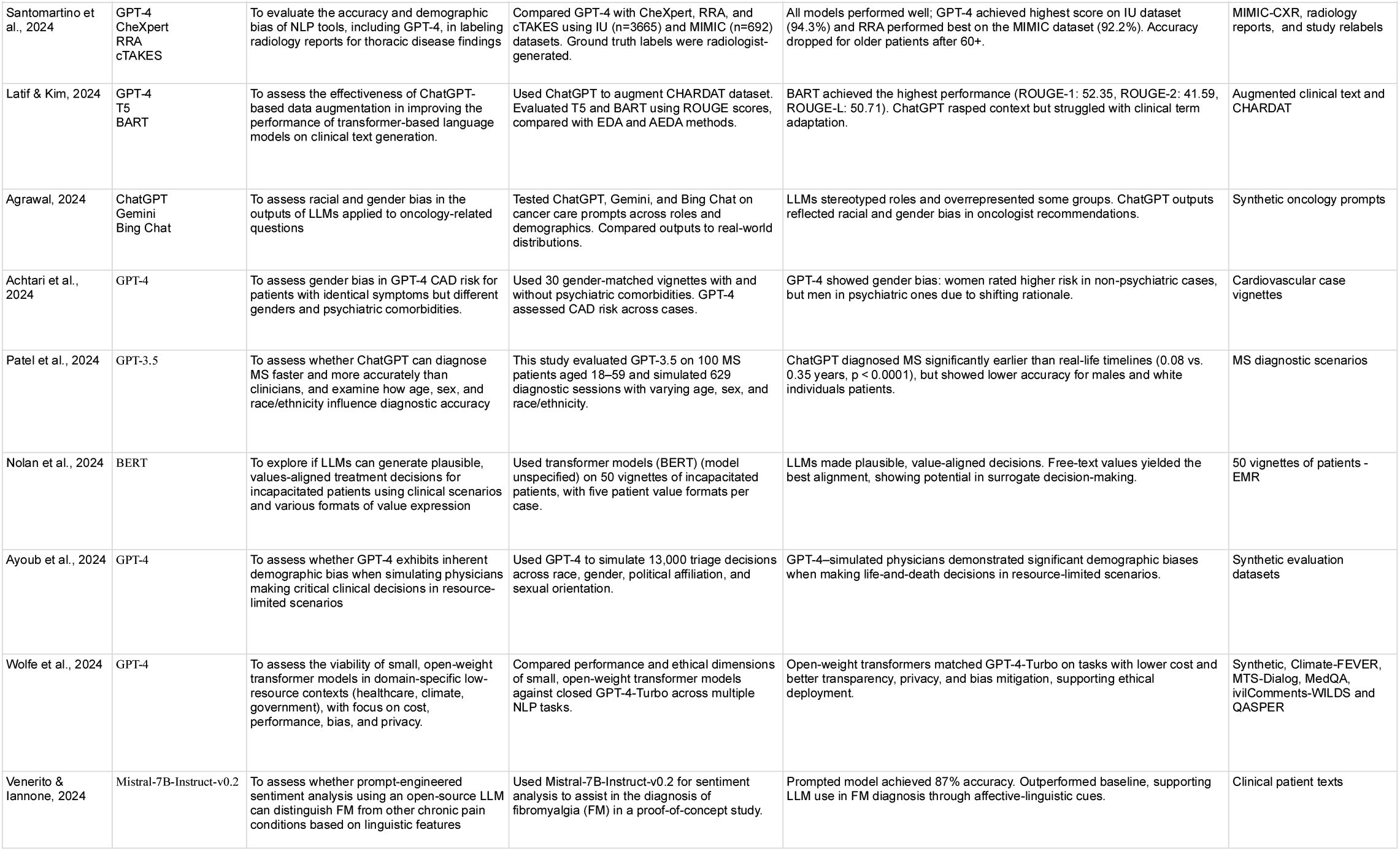
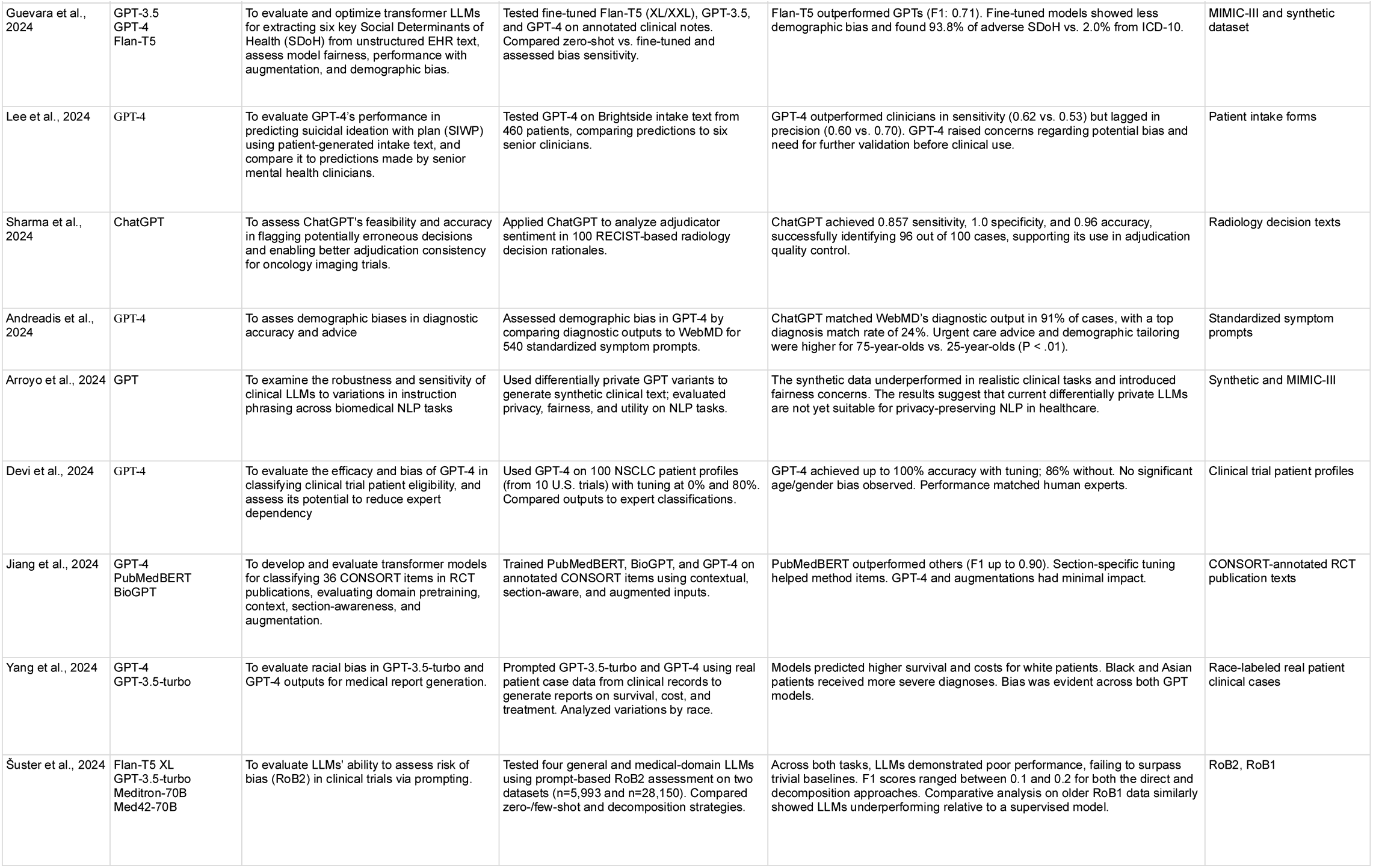
Full database search strategy used for the scoping review.

## Data Availability

All 56 articles analyzed during this study, and the data availability table, are available on OSF and can be accessed via https://osf.io/t6y48/.

## Code availability

There are no codes developed in this study.

## Supplementary Material

Table 1 provides a comprehensive summary of all fifty-six studies included in this scoping review, outlining for each article the authors, year of publication, model architecture, study objective, methodological approach, key findings, and dataset type. This table serves as the empirical foundation for the review, offering a transparent account of how transformer-based NLP models have been applied within healthcare and how equity-related considerations have been addressed across diverse contexts. The extracted data align with the PRISMA-ScR and Arksey and O’Malley frameworks, ensuring systematic and reproducible synthesis. The table captures substantial variation in research scope and design, illustrating that algorithmic equity remains the most frequently explored dimension, primarily through fairness audits, subgroup performance analyses, and bias-mitigation strategies in clinical text classification and diagnostic prediction tasks. Data diversity and representativeness appear less consistently, with most studies relying on English-language or single-institution datasets, revealing persistent limitations in linguistic and demographic inclusivity that we conceptualize as data diversity debt. Participatory and inclusive design is rarely documented, and where present, engagement is typically confined to clinician validation or expert review, with minimal involvement of patients, caregivers, or community stakeholders. The table highlights both the growing sophistication of transformer applications and the urgent need to embed equity and inclusivity throughout the entire model lifecycle—from data collection and training to evaluation and real-world deployment.

## Acknowledgements

The research was undertaken thanks in part to funding from the Connected Minds Program, supported by the Canada First Research Excellence Fund, Grant #CFREF-2022-00010. Also, Resources used in preparing this research were provided, in part, by the Province of Ontario, the Government of Canada through CIFAR, and companies sponsoring the Vector Institute. Elham Dolatabadi’s research was supported by a Natural Sciences and Engineering Research Council of Canada (NSERC) Discovery Grant and a Canadian Institutes of Health Research (CIHR) Special Call from the Center for Research on Pandemic Preparedness and Health Emergencies.

